# Possible Role of P-selectin Adhesion in Long-COVID: A Comparative Analysis of a Long-COVID Case Versus an Asymptomatic Post-COVID Case

**DOI:** 10.1101/2022.03.09.22271297

**Authors:** Michael Tarasev, Sabrina Mota, Xiufeng Gao, Marta Ferranti, Aliya U. Zaidi, Bryan Hannan, Patrick Hines

**Author notes:** Correspondence to: Michael Tarasev, Functional Fluidics, 440 Burroughs St., Ste. 520, Detroit, MI, USA, (email).

## Abstract

**Background:** Long-term outcomes of severe acute respiratory syndrome coronavirus 2 (SARS-CoV-2) are now recognized as an emerging public health challenge - a condition termed Long-COVID. The pathophysiology of Long-COVID remains to be established. Functional P-selectin activity, implicated in COVID-19 sequalae, was measured between two convalescent COVID-19 subjects, one with (Long-COVID subject) and another without Long-COVID symptoms.

**Methods:** Flow adhesion of whole blood or isolated white blood cells to P-selectin (FA-WB-Psel and FA-WBC-Psel) was measured using a standardized microfluidics clinical assay; impedance aggregometry with a collagen agonist was measured using model 590 Chrono-Log impedance aggregometer; standard laboratory assays were performed to evaluate changes in blood chemistries.

**Results:** For both subjects, hemoglobin, WBC, platelet counts, electrolytes and ferritin were within normal reference ranges, with FA-WB-Psel significantly elevated compared to healthy controls (p< 0.01). *In vitro* treatment of whole blood samples with crizanlizumab (anti-p-selectin monoclonal antibody) within the clinical dose range (10 μg/ml) mL) inhibited FA-WB-Psel only in samples from asymptomatic post-COVID subject, with the Long-COVID subject sample requiring close to 5-fold elevated dose to achieve a response. Pronounced inhibition of P-selectin adhesion of isolated leukocytes was observed for both subjects in autologous platelet-poor plasma and buffer. Impedance aggregometry showed greater baseline platelet aggregation to collagen in the Long-COVID sample, although both samples responded similarly to aspirin-induced platelet inhibition.

**Conclusions:** Presented results suggest that elevated platelet activation in Long-COVID subject may be associated with increased P-Selectin activity. The results are discussed in terms of possible use on P-selectin inhibition therapies in treating Long-COVID.

## Background

Severe acute respiratory syndrome coronavirus 2 (SARS-CoV-2) causes the Coronavirus disease 2019 (COVID-19), which is characterized by severe vascular complications associated with endothelial dysfunction and overproduction of inflammatory mediators, with pathology extending to the cellular level inducing a pro-coagulant state. Research efforts have focused on the acute phase of the disease; however, long-term outcomes for people recovering from COVID-19 are now recognized as an emerging public health challenge. Although most people show complete recovery within a few weeks, some continue to experience a range of symptoms long after their initial recovery which is often referred to as “Long-COVID” or post-acute sequelae of COVID-19 (PASC). While the definition of Long-COVID is still evolving, it has been suggested to include persistence of symptoms or development of related pathologies beyond 3-4 weeks from the onset of acute symptoms of COVID-19 [1-3]. Specifically, it was proposed that at least 2 periods of illness appear to be contributing to morbidity beyond acute SARS-CoV-2 infection: a post-acute hyperinflammatory illness and late inflammatory and virological sequelae. The post-acute hyperinflammatory condition manifests approximately 2 to 5 weeks after the onset of SARS-CoV-2 infection and is characterized by systemic inflammation that can occur in organs distinct from those directly affected by the virus, after host clearance of the SARS-CoV-2 infection. The pathophysiology of this multisystem inflammatory syndrome likely reflects a dysregulated host immune response [4].

The late sequelae or Long-COVID includes a wide range of cardiovascular, pulmonary, neurological, and other physiological manifestations [5]. Typically, patients with Long-COVID report different combinations of symptoms: fatigue, difficulty thinking or concentrating (sometimes called “brain fog”), headache and migraines, loss of smell or taste, hypertension, dizziness on standing, heart palpitations, chest pain, difficulty breathing or shortness of breath, cough, joint or muscle pain, depression or anxiety, hair loss, fever, symptoms that get worse after physical or mental activities. Symptoms attributed to systemic inflammation and impaired microvascular blood flow are of particular concern. These symptoms can continue for an undetermined duration, and it remains unknown to what extent these symptoms can evolve into chronic or irreversible health conditions. There is limited information about the underlying pathophysiology, disease duration, or long-term prognosis of persons affected by Long-COVID. It seems likely that individual patients with Long-COVID would manifest the symptoms through different biological drivers with suggested contributions from viral-induced injury to one or multiple organs, persistent reservoirs of SARS-CoV-2 in certain tissues, re-activation of pre-existing pathogens stimulated by COVID-19 immune dysregulation, SARS-CoV-2 interactions with host microbiome/virome, ongoing activity of primed immune cells, and escalating autoimmunity among other possibilities [3].

The interest in Long-COVID continues to increase as new reports detailing the condition continue to emerge. A number of investigators reported different prevalence of persisting complications in patients recovered from COVID-19. A study published in JAMA presented cardiac magnetic resonance imaging data showing 78% patients exhibiting cardiac involvement and 60% having an ongoing myocardial inflammation [6]. It was also reported that two-months post the infection among the adults with non-critical COVID-19, two thirds had complaints including severe dyspnea or asthenia, chest pain, palpitations, headache, myalgia, and fever [7], while a study published in BMJ reported about 10% of those who had COVID-19 experienced symptoms beyond three weeks, with an even smaller proportion experiencing them for months [2].

A recent FAIR Health study reviewed a total of 1,959,982 COVID-19 patients for the prevalence of Long-COVID condition 30 days or more after their initial diagnosis with COVID-19 and reported that about 23% had at least one Long-COVID symptom [8]. The five most common Long-COVID conditions across all ages, in order from most to least common, were pain, breathing difficulties, hyperlipidemia, malaise/fatigue, and hypertension. Long-COVID conditions were found to a greater extent in patients who had more severe cases of COVID-19, but also in a substantial share of asymptomatic patients. Of patients who were hospitalized with COVID-19, the percentage that had a Long-COVID condition was 50%, while for patients who were symptomatic but not hospitalized, 27.5%; and 19% for patients who were asymptomatic for COVID-19 [1]. Without a clear clinical picture of the underlying mechanisms, the treatment of Long-COVID syndrome is mainly limited to supportive care and symptomatic control, and its long-term impacts on quality of life of COVID-19 survivors remains unclear.

Commonly reported for COVID-19 positive patients are lymphocytopenia, as well as elevated levels of D-dimer, lactate dehydrogenase, ferritin, Von Willebrand factor, Factor VIII, and inflammatory markers like C-reactive protein, IL-6 and erythrocyte sedimentation rate [9-12]. Despite seeming trends, the reports do not present consistent picture of COVID-19-associated changes and can be hard to reconcile with existing patient-to-patient variability as the reportedly altered values in many cases remain within the normal physiological range. In other cases, both elevated and depleted biomarker values had been observed as is e.g., the case of platelet counts, where both thrombocytopenia and thrombocytosis have been reported [12]. The analysis is further complicated by biomarker significant dependence on disease severity and associated co-morbidities, as well as on wide range of possible patient clinical trajectories defined by an interplay of immunological, inflammatory, and coagulative processes.

COVID-19 association with platelet activation is well documented [13]. Typically, through binding to PSGL-1 on WBC, P-selectin overexpressed on platelets would enhance platelet aggregation and thrombus formation. Indeed, viral-induced coagulopathies including arterial thrombotic events like stroke and ischemic limbs as well as microvascular thrombotic disorders have been observed in SARS-CoV-2 infection with platelet hyperactivity suggested as the key element in disease thrombotic manifestations [14]. Clinically, in patients hospitalized with COVID-19, platelet hyperactivity had been associated with adverse events including thrombosis and death [15]. Such events are often, but not always, correlated with elevation of thrombosis biomarkers like D-dimer and fibrin/fibrinogen-degradation products. Interestingly, some studies did not observe a correlation between levels of D-dimers and platelet activation [16].

Inflammatory and coagulative sequelae are mediated by selectins including P, E, and L selectin and cell adhesion molecules (ICAM-1, VCAM-1), with P-Selectin in particular gaining attention. Located within the α-granules of platelets and the Weibel-Palade bodies of endothelial cells, P-Selectin rapidly translocate to the cell surface following activation. P-selectin in platelets interacts with P-Selectin glycoprotein ligand-1 (PSGL-1) on leukocytes promoting platelet-leukocyte aggregate formation, release of procoagulant microparticles, and upregulation of a several leukocyte cytokines. It is also involved in platelet-platelet aggregation, which is major factor in arterial thrombosis [17]. Most investigators report elevated levels of P-Selectin in SARS-CoV-2 positive patients, with seemingly larger elevation of P-selectin in more severe disease states. However, a number of studies also report comparable levels of P-selectin in COVID-19 patients and healthy controls [18].

Studies also indicated elevated soluble P-selectin (sP-selectin) levels in plasma of COVID-19 patients [19], with such levels suggested as an early marker of thromboembolism [20]. sP-selectin predominantly originate through proteolytic cleavage of the transmembrane protein (shedding). For example, activated platelets can shed P-Selectin in hours after activation [21]. The process is mediated through leukocyte P-Selectin glycoprotein ligand-1 (PSGL-1) interaction with transmembrane P-Selectin on platelets or endothelial cells [22]. However, despite its elevated plasma concentration, mostly monomeric sP-selectin, by itself is expected to have limited impact on P-Selectin-associated pathology, as its prior dimerization is likely required to promote inflammation and coagulation [22].

While some understanding seems to emerge regarding the active phase of COVID-19, the pathophysiology involved in persisting Long-COVID remain essentially unknown. Part of the reason is that long-term follow-up presently is too limited to reveal the full scope of all potential COVID-19 consequences. Another is the natural focus of the research community on understanding and combatting of the active phase of the infection. A recent review presents the multitude of mechanisms potentially involved in the development of Long-COVID symptoms [23]. Diversity of the proposed mechanisms underlines both the complexity of Long-COVID pathology and the need for further investigation on the resolution of this new health problem. Presently we report changes in platelet reactivity, blood cell adhesion on P-Selectin and response to *in-vitro* P-Selectin inhibition therapy in subjects after SARS-CoV-2 infection with and without severe Long-COVID symptoms.

## Methods

### Study Subjects and Sample Collection

Two convalescent COVID-19 subjects, one with Long-COVID symptoms (LC) and the other asymptomatic post-COVID (PC) were recruited according to the protocols FF-RBC-001 and FF-RBC-003v2 approved by Institutional Review Board of the Institute for Regenerative and Cellular Medicine. Throughout this paper, the two subjects will be referred to as, the Long-COVID subject ,(LC), and the asymptomatic post-COVID subject (PC). The blood draws were obtained via venipuncture after a 12-h fast. Whole blood was collected into EDTA tube for CBC, aggregometry, and flow adhesion assays, 3.2% sodium citrate tube for D-dimer, and serum separation tubes for interleukins, COVID-19 antibodies and for calcium, LDH, iron, TIBC and ferritin panel that were transferred to Quest Diagnostic Lab for analysis. Reference ranges are those provided by the laboratory unless specified otherwise. Both subjects consented to take part in the present study and for the results to be published.

### Flow Adhesion Assays

Flow adhesion of whole blood to P-Selectin (FA-WB-Psel) and Flow adhesion of white blood cells to P-Selectin (FA-WBC-Psel) were conducted as described previously [24]. Briefly, isolated white blood cell (i-WBC) suspensions were prepared according to a standardized protocol (HetaSep™, StemCell Technologies). Whole blood samples (1:1 diluted with HBSS buffer) or i-WBC (5× 10^6^ cells/mL) were perfused through P-Selectin-coated microfluidic channels at 1 dynes/cm^2^ for 10 or 6 minutes respectively, washed to eliminate non-adhering cells, with resultant adhesion quantified manually by an independent trained observer to generate an adhesion index (cells/mm²) using a previously described protocol [25]. For drug treatment conditions, samples were incubated for 5 minutes with crizanlizumab at the final concentrations as required before assessment by the flow adhesion assay.

### Impedance Aggregometry

Platelet aggregation in whole blood samples was tested with an impedance aggregometer (Model 590; Chrono-Log Corporation, Havertown, PA) and analyzed using Aggro/Link®. Whole blood samples were collected into 3.2% sodium citrate tubes, transferred at room temperature, and tested within 4 hours after the blood draw. The sample was split into control (1% DMSO treated) and aspirin treatment (100µM). Briefly, after a stable baseline (steady state) had been established, the agonist collagen (Chrono-Par, Chrono-Log Corporation, Havertown, PA) was added to the sample to a final concentration of 2µg/mL, and the aggregation was monitored for 10 minutes. Instrument normal control reference values (Table 3) are for normal donor with no previous exposure to COVID19 tested same as post-COVID subjects. Note, that sample collection in citrate tubes required for other assays could have resulted in lower overall values than would have been observed in EDTA [26].

### Statistical Analysis

The data is presented as mean ± standard deviation (mean ± SD) with Student t-test paired or non-paired as appropriate used for assessment of statistical significance of the differences. Results were deemed significant for comparisons where two-tailed *p* < .05.

## Results

### Clinical Sequelae

#### Post-COVID subject with severe Long-COVID symptoms

LC, is a 26-30 year old female with no history of chronic medical conditions, was not on any prescription medication, and had no prior health complaints. She was diagnosed with SARS-CoV-2 by PCR and presented with COVID-19 symptoms one week following a known exposure, including coughing, fever, difficulty breathing, cough, progressively worsening fatigue, brain fog, loss of taste and smell, and headache. In LC’s own words, “*I had severe issues breathing unless I was laying down in a certain position. When I did need to sit or stand, I would have a coughing attack barely being able to breathe or be severely short of breath. My throat was extremely irritated. I was disoriented for about 3-4 days. I lost my taste and smell before the major symptoms presented but regained most of both shortly after I began to recover. I had a constant burning in my nose, almost like I had to sneeze. Headaches were nearly unbearable and remained consistent for weeks afterward, with no headache medication able to alleviate*.” LC was not hospitalized, managing symptoms at home. At most severe (10-12 days after the infection), she was on bed rest, not responsive, refusing food or drink. The symptoms started to alleviate when the fever was broken and gradually receded over the course of another week. After the resolution of the infection, LC experienced a wide range of symptoms (Table 1) typical for those reported in Long-COVID cases. While some symptoms started to alleviate to different degree over time, many of them remain quite severe even 5 months after the infection (Table 1).

**Table 1.** Subject (LC) Long-COVID symptoms development

| Symptom<sup>1</sup> | Time after SARS-CoV-2 exposure, weeks |  |  | Comment |
| --- | --- | --- | --- | --- |
|  | 11 | 20 | 28 |  |
| Brain fog | 8 | 8 | 6 | Frequent random moments of forgetfulness and fog that started only after the recovery from SARS-CoV-2 infection |
| Headaches | 7 | 6 | 5 | Ongoing dull headaches: by 28 weeks after the exposure decreased in frequency to 2-4 times a week, but not responding to treatment by normal headache medication |
| Fatigue | 8 | 8 | 7 | Subject commented that at 28 weeks “Fatigue has begun to wear off, but still hits hard at different times” |
| Hair Loss | 9 | 8 | 1 | Was ongoing, but stopped at about around 24 weeks |
| Burning in the nose | 8 | 7 | 3 | Added and highlighted by the subject. Described as an ongoing sensation ‘burning in the nose like I have to sneeze” |
| Menstrual cycle | +++ | ++ | + | Cycle regular during the active infection with a 3-weeks additional delay after the recovery. About 5 weeks cycle afterwards (+++) slowly moving back to normal after 3 months post exposure (++) , normalizing at about 28 weeks after the infection |

<sup>1</sup>Self-reported severity of symptoms is shown with 0 being barely ever and 10 being very bad/frequent

#### Post-COVID subject with no Long-COVID symptoms

PC, is a 16-20 year old male, with no history of chronic medical conditions. He was diagnosed with COVID-19 and presented mild symptoms during the active infection lasting approximately 3-4 days. He was asymptomatic three weeks before participating in the study and remained asymptomatic 20 weeks later. He achieved full recovery with no lingering Long-COVID symptoms present at any time since infection resolution.

### Clinical Chemistry

#### LC

Despite severe Long-COVID symptoms, clinical chemistry results of the Long-COVID subject (LC) show little deviation from the normal ranges (Table 2). As an exception, at 20 weeks after the infection with SARS-CoV-2, the subject shows slightly elevated RBC count (5.26*10^12^ cells/L), as compared to the normal reference range of 3.8 - 5.1*10^12^ cells/L. At normal blood hemoglobin value, this slight increase in RBC count is correlated with mean cell hemoglobin concentration (MCH) at the lower end of the normal range. In parallel, metabolic panel shows a decreased total iron and Transferrin-iron saturation percentage values on the background of normal Total Iron Binding Capacity (TIBC), that could be functionally related to increased RBC counts. The values for Ferritin and LDH, often reported as elevated in COVID-19 patients, as well as bilirubin, were within the normal reference range, with the subject thus presenting no evidence of extraordinary hemolysis that could be accompanying SARS-CoV-2 infection [12, 27]. D-Dimer value was elevated compared to the reference range (1.02 vs normal value of <0.5 mcg/mL). An increase in D-Dimer associated with increased in thrombosis risk has been well documented in SARS-CoV-2 infection [28] with significant, 3-4-fold rise in D-dimer levels (to 1.2 ± 1.4 mcg/mL) in the early stages of COVID-19 linked to poor prognosis. All other values are within the normal reference ranges at both 11-and 20-weeks post infection.

**Table 2.**

| | Post-COVID<br>subject (male) | Long-COVID subject (female) | | | Average reported<br>values (mean ± SD) |
| --- | --- | --- | --- | --- | --- |
| Weeks past the resolution of SARS-CoV-2 infection | 2.5 weeks | 11 weeks | 20 weeks | Reference range<sup>1</sup> | During active COVID-19 |
| Complete blood count (CBC) |  |  |  |  |  |
| WBC, bil/L | 7.7 | 7.8 | 9.9 | 3.3 – 10.7 | 6.0 ± 2.0 |
| RBC, tril/L | 5.07 | 4.92 | ↑ 5.26 | 3.8 - 5.1 | 4.2 ± 0.4 |
| Hemoglobin, g/dL | 14.6 | 13.4 | 14.2 | 12.1 - 15.0 | 13 ± 1.1 |
| Hematocrit, % | 43.6% | 41.5% | 43.6% | 35.4% - 44.2% | 41.6% ± 2.7% |
| MCV, fL | 86.0 | 84 | 82.9 | 80 - 100 | 91.5 ± 5.2 |
| MCH, pg | 28.8 | 27 | 27 | 27 - 33 | 30.6 ± 2.1 |
| MCHC, g/dL | 33.5 | 32 | 32.6 | 32.0 - 35.0 | 33.3 ± 1.1 |
| RDW-CV, % | 11.7% | 14% | 13% | 11% - 15% | 14.3% ± 1.7% |
| Platelets, bil/L | 264 | 318 | 310 | 150 - 400 | 183 ± 39 |
| MPV, fL | ↑ 12.8 | N/A | 10.2 | 7.5 - 12.5 | 7.8 ± 4.2 |
| Metabolic Panel and electrolytes |  |  |  |  |  |
| Sodium, mmol/L | 144 | 140 | N/A | 135 - 145 | 138.7 ± 2.4 |
| Potassium, mmol/L | 4.3 | 4.7 | N/A | 3.6 - 5.2 | 4.2 ± 0.5 |
| Chloride, mmol/L | ↓ 97 | 104 | N/A | 98 - 111 | 102.8 ± 3.1 |
| Carbon Dioxide (CO<sub>2</sub>), mmol/L | ↓ 16 | 23 | N/A | 20 - 29 | ↓ 12 ± 12.3 |
| Ferritin, ng/mL | 134 | N/A | 26 | 11-172 | ↑↑ 1018 ± 533 |
| Iron, Total, mcg/dL | 163 | N/A | ↓ 35 | 40 - 190 | 150 ± 55 |
| Iron Binding Capacity (TIBC), mcg/dL | 394 | N/A | 358 | 250 - 450 | 390 ± 53 |
| Transferrin-iron saturation percentage (TSAT), % | 41% | N/A | ↓ 10% | 16% - 45% | 29% ± 10% |
| Protein Total, g/dL | 7.7 | 7.1 | N/A | 6.4 - 8.3 | 6.8 ± 0.7 |
| Albumin, g/dL | 4.8 | 4.4 | N/A | 3.5 – 5.1 | 3.9 ± 0.5 |
| Globulin, g/dL | 2.9 | 2.7 | N/A | 2.2 – 4.0 | 2.7 ± 0.3 |
| Alkaline Phosphatase, U/L | 79 | 82 | N/A | 33 - 120 | 84 ± 49.3 |
| Aspartate Aminotransferase (AST), U/L | 25 | 22 | N/A | < 35 | ↑ 44.2 ± 43.0 |
| Alanine Aminotransferase (ALT), U/L | 15 | 24 | N/A | 8 - 37 | ↑ 42.3 ± 34.9 |
| Bilirubin Total, mg/dL | ↑ 1.3 | 0.6 | N/A | 0.3 - 1.2 | ↑ 1.3 ± 1.0 |
| Other parameters |  |  |  |  |  |
| D-Dimer, mcg/mL FEU | <0.19 | N/A | ↑↑ 1.02 | <0.50 | ↑↑ 1.2 ± 1.4 |
| Total LDH, U/L | ↑ 278 | N/A | 180 | 100-200 | ↑↑ 370 ± 344 |

Values above the normal reference range are indicated by ↑ for moderate increase and by ↑↑ for large increase, and values below the normal reference range indicated by ↓ for moderate increase and
by ▼▼ for large increase. All other values are within the reference range. Abbreviations: WBC, white blood cell; RBC, red blood cell; MCV, mean corpuscular volume; MCH, mean corpuscular hemoglobin; MCHC, mean corpuscular hemoglobin concentration; RDW-CV, red cell distribution width; MPV, mean platelets volume.
<sup>1</sup> The reference ranges are those provided by Quest Diagnostics.

#### PC

Clinical chemistry parameters were within the normal reference ranges for the asymptomatic post-COVID subject (PC), with the exception of slightly elevated total bilirubin (1.3mg/dL, compared to the reference range 0.1-1.2 mg/dL) and total LDH (278 U/l, reference range 110-230 U/L). Simultaneous elevation of LDH and bilirubin may be indicative of slightly elevated rate of hemolysis on the background of compensating erythropoietic activity and thus with no impact on RBC count or total hemoglobin values. That supposition is supported by the concurrently decreased Red Blood Cell Distribution Width (RDW), with such a decrease being potentially associated with decreased average RBC lifespan in circulation resulting in smaller contribution of older and lower volume cells to RDW [29].

Interleukin 1 Beta (IL1β) and interleukin 4 (IL4) were in the normal reference ranges for both subjects (Table 2).

### Flow Adhesion on P-Selectin

#### Whole Blood

Flow adhesion of whole blood on P-Selectin (FA-WB-PSel) was significantly elevated for both Long-COVID (LC and asymptomatic post-COVID (PC) subjects (590 ± 260 and 1,100 ± 250 cells/mm^2^ correspondingly) as compared to normal subjects (typically, 50 or less cells/mm^2^ if with no significant inflammation processes).

#### LC

Measurements of flow adhesion of whole blood in the presence of crizanlizumab, a monoclonal antibody against P-selectin, resulted in a dose-dependent decline of adhesion to P-selectin for the asymptomatic post-COVID subject’s sample. Such crizanlizumab-induced inhibition of adhesion to P-selectin reached about 60 percent at 10 µg/mL. This dose correlates to the reported mean steady-state serum crizanlizumab concentration at 10.5 to 15.0 μg/mL for high drug administration dose (5 mg/kg) [30]. At 60 µg/mL of crizanlizumab, 90% of inhibition of FA-WB-Psel was reached (Figure 1A).

**Figure 1.**
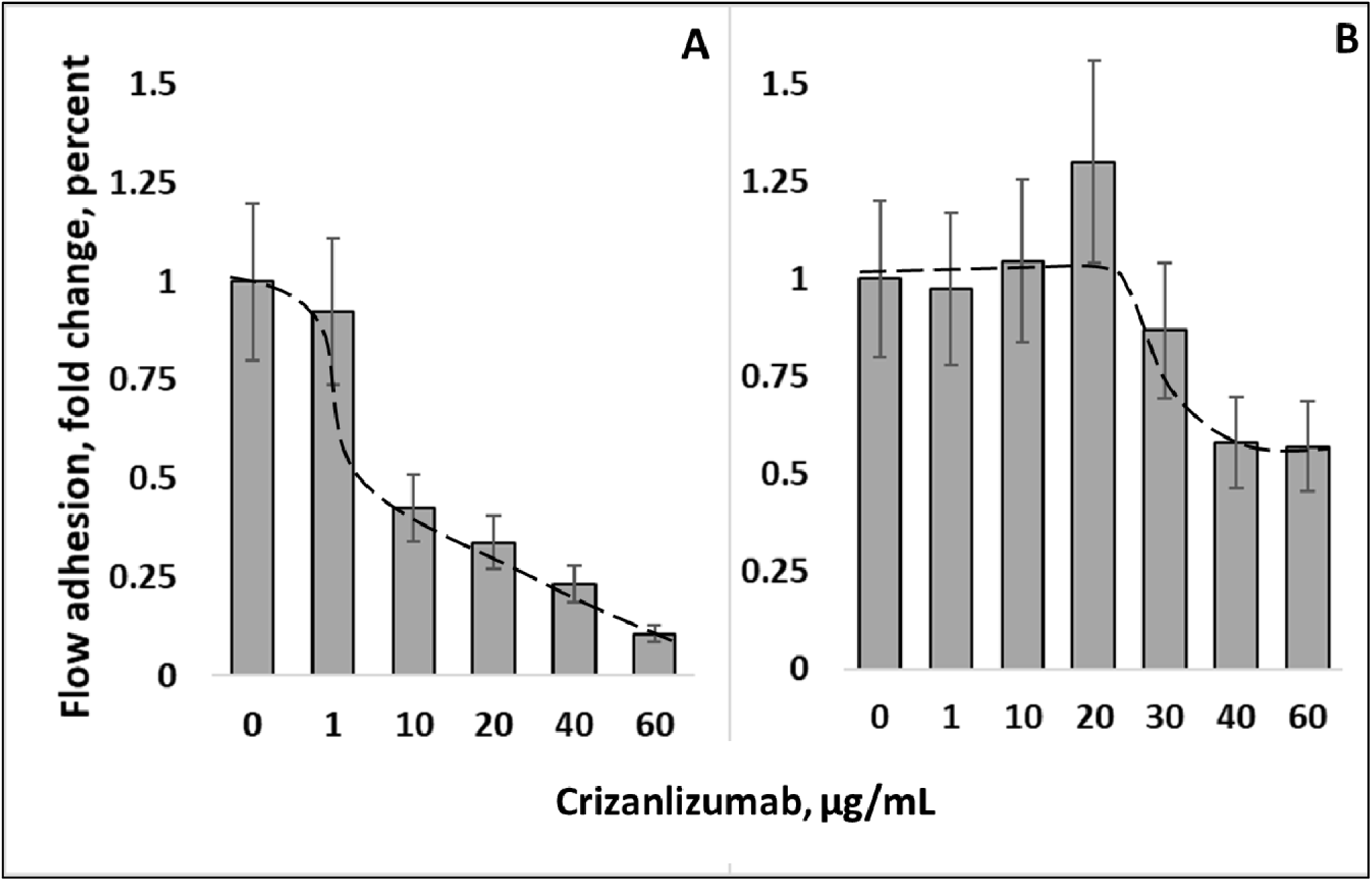
Changes in whole blood flow adhesion on P-Selectin substrate in the presence of crizanlizumab. A. Asymptomatic post-COVID (PC) and B. Long-COVID (LC) subjects. Trend lines are for an illustrative purpose only. Adhesion is shown as a fold change relative to the no-drug baseline.

#### PC

For the Long-COVID subject with severe symptoms, supplementation of whole blood with crizanlizumab in doses up to 20 μg/mL range did not result in any measurable inhibition of cell adhesion to P-selectin for the symptomatic patient sample. At the elevated drug doses (50-60 µg/mL) up to 50% inhibition of adhesion P-selectin was observed, which is about half of that recorded with the asymptomatic subject at the same dose (Figure 1B).

### Isolated White Blood Cells in buffer

#### PC&LC

Flow adhesion of isolated White Blood Cells (i-WBC) to P-selectin (FA-WBC-Psel) was similar between Long-COVID and asymptomatic post-COVID subjects, and in HBSS buffer was 1490 ± 30 and 1590 ± 110 cells/mm^2^ correspondingly. When i-WBCs were suspended in autologous plasma (see Material and Methods), adhesion on P-selectin was decreased by 4.2- and 3.8-fold to 350 ± 20 and 420 ± 50 cells/mm^2^ for asymptomatic post-COVID and Long-COVID subjects correspondingly (Figure 2).

**Figure 2.**
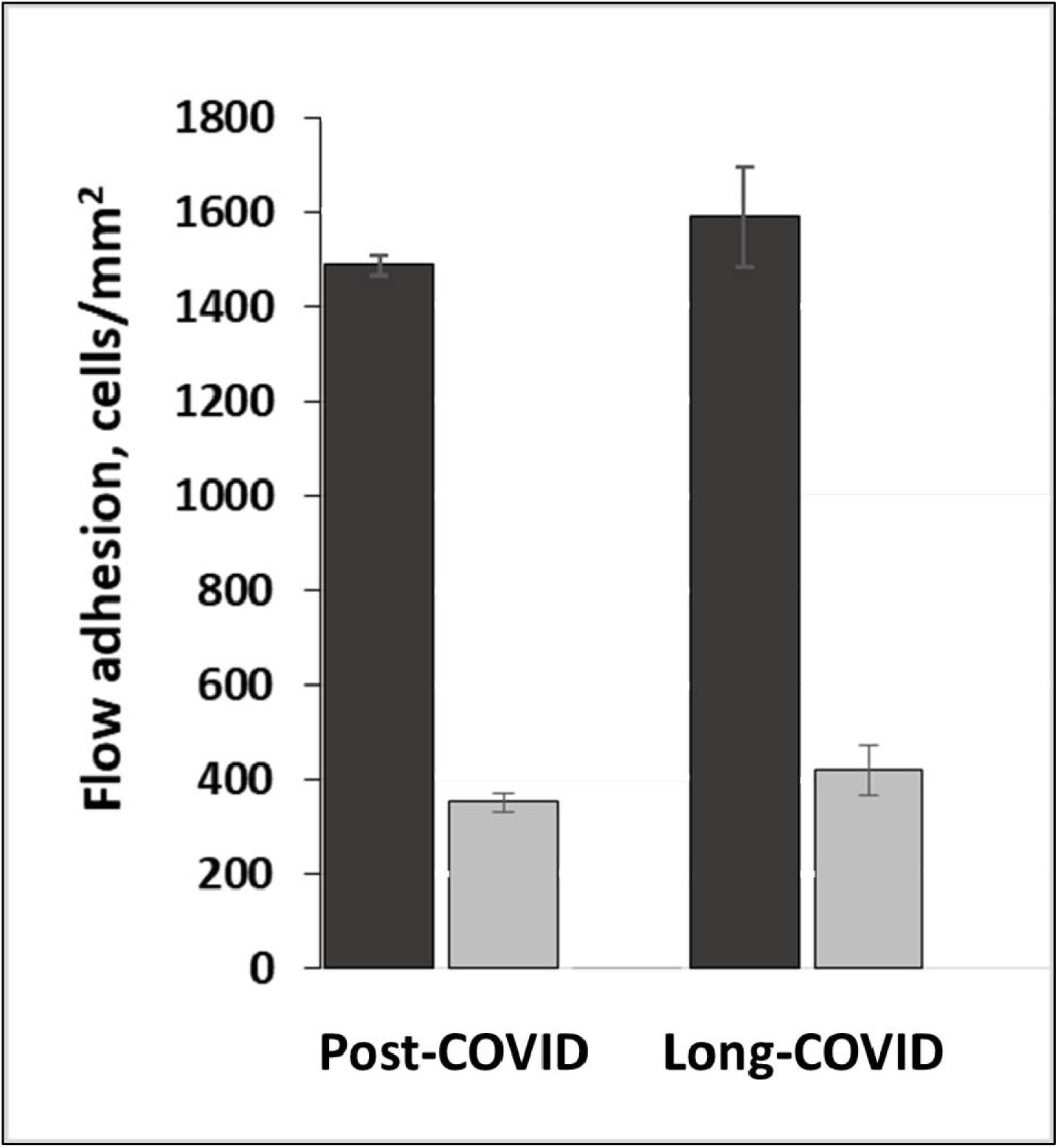
Isolated white blood cell flow adhesion on P-Selectin substrate for asymptomatic post-COVID (PC) and Long-COVID (LC) subjects. Adhesion in HBSS buffer (▪) and in patient’s own platelet poor plasma 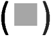.

#### PC

For the asymptomatic post-COVID subject, supplementation of buffer-suspended i-WBC with crizanlizumab resulted in a dose-dependent inhibition of WBC adhesion with the magnitude of inhibition similar to that observed in Whole Blood (the difference lacked statistical significance).

#### LC

For the subject with Long-COVID symptoms, supplementation of i-WBC with crizanlizumab showed nearly complete (about 95%) inhibition of cell adhesion to P-Selectin. Such un-expectantly strong adhesion inhibition had been observed event at 1 µg/mL dose of crizanlizumab, which is ten times lower than the standard clinical drug dose (Figure 3).

**Figure 3.**
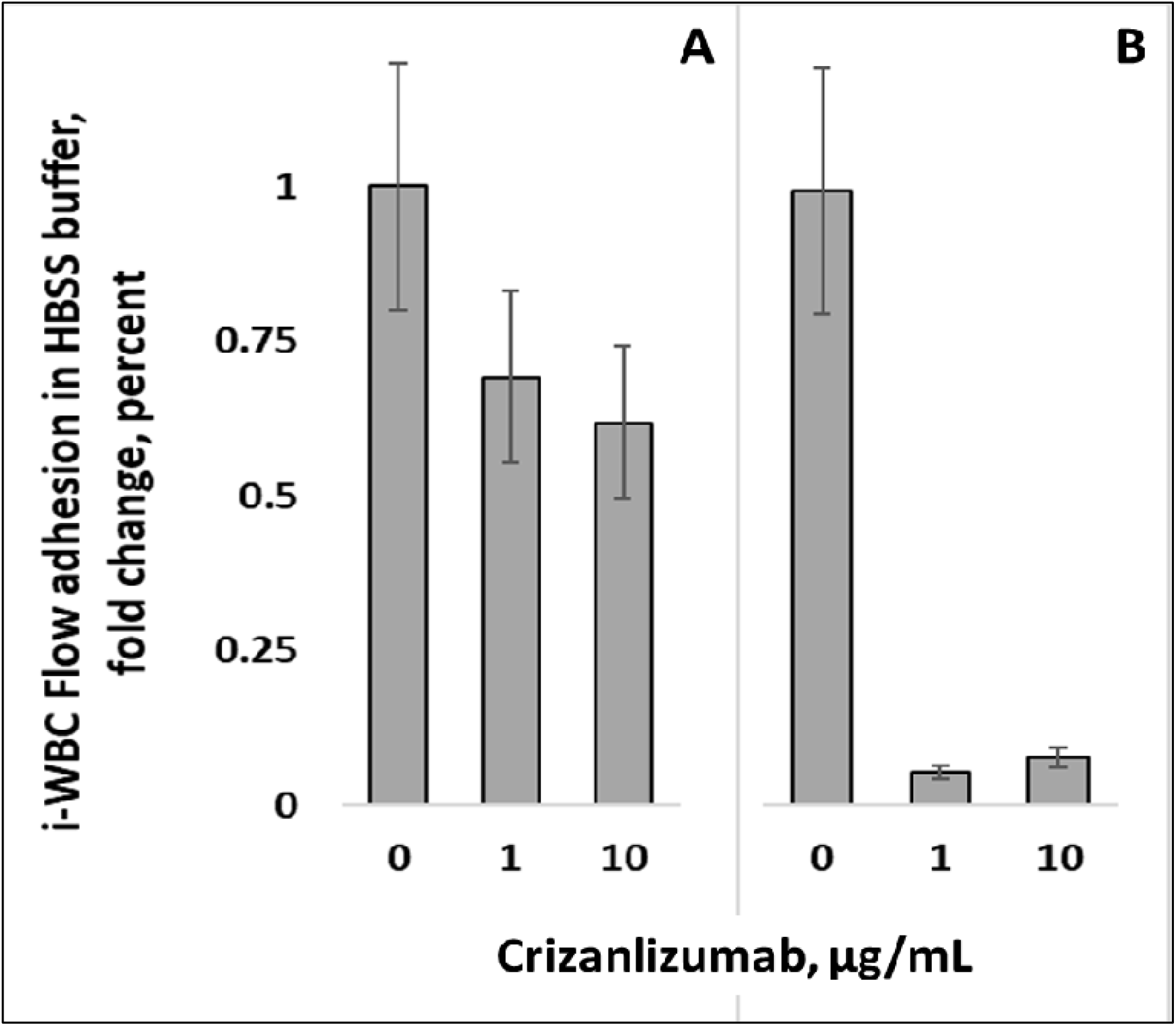
Flow adhesion on P-Selectin substrate of isolated WBC in HBSS buffer (see Materials and Methods) supplemented with crizanlizumab for asymptomatic post-COVID (A) and Long-COVID (B) subjects. Adhesion is shown as a fold change relative to the no-drug baseline.

### Isolated White Blood Cells in autologous Platelet Poor Plasma

#### PC&LC

Crizanlizumab supplementation of same i-WBC suspended in autologous Platelet Poor Plasma (PPP) resulted in no significant inhibition at sub-clinical crizanlizumab dose corresponding to 1 µg/mL plasma concentration and in pronounced inhibition of adhesion to P-selectin (about 80 % for asymptomatic and 65 % for symptomatic subjects, Figure 4). Similar results with adhesion inhibited by about 70-80 % were obtained when crizanlizumab-induced inhibition of adhesion was measured on i-WBC in blood-type complimentary platelet poor plasma from healthy subjects (data not shown). For the Long-COVID subject similar results had been obtained on samples collected at 11 and 20 weeks after the infection (at approximately 8 and 16 weeks after the resolution of acute COVID-19).

**Figure 4.**
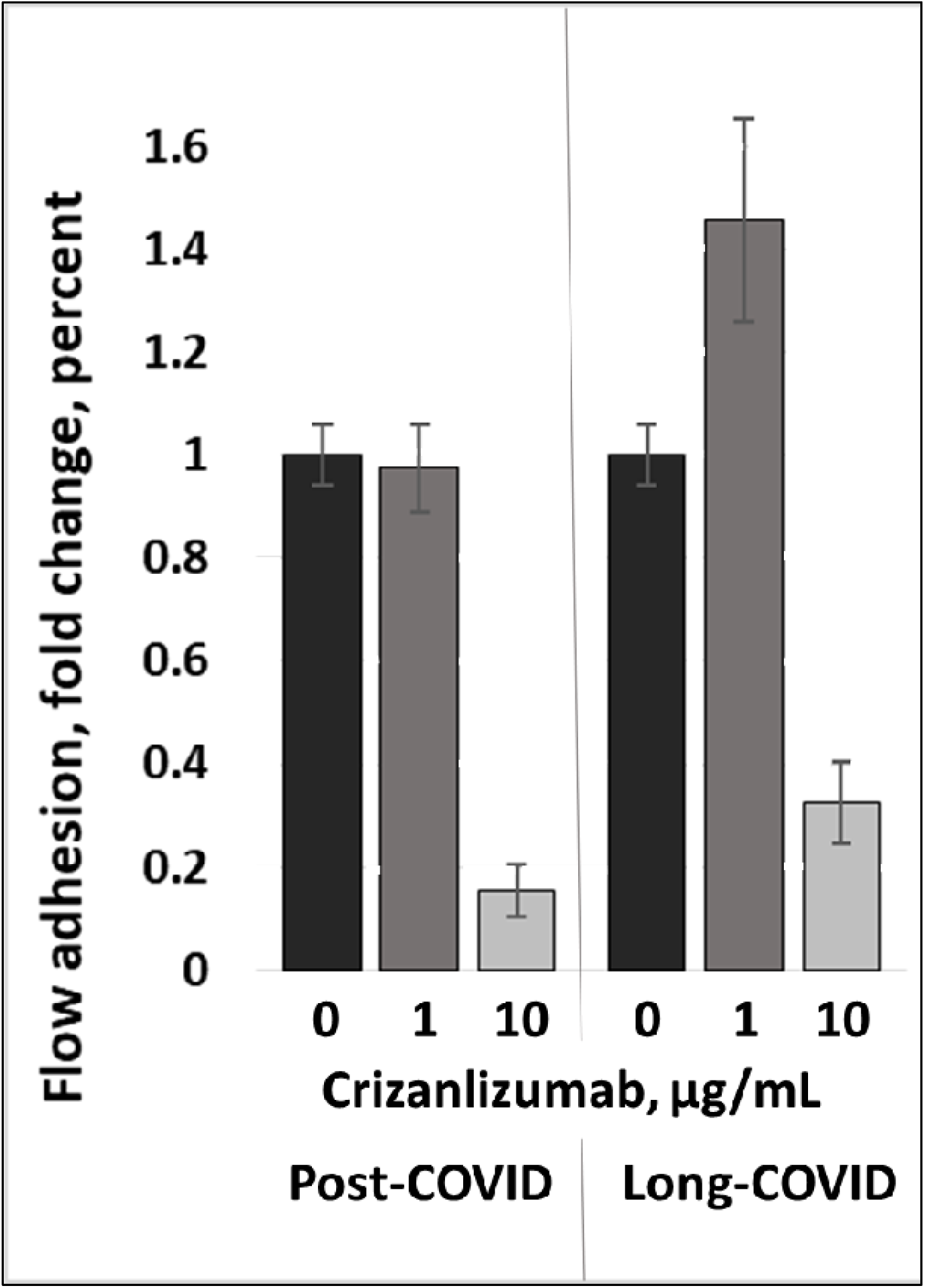
Flow adhesion on P-Selectin substrate of isolated WBC in patient’s own platelet poor plasma for asymptomatic post-COVID and Long-COVID subjects in the presence of crizanlizumab. Adhesion is shown as a fold change relative to the no-drug baseline.

### Whole Blood Aggregometry

#### PC&LC

Whole blood impedance aggregometry showed that maximum amplitude, area under the curve (AUC) and maximum rate were all elevated in post-COVID subjects, both with Long-COVID and asymptomatic, as compared to normal donors (see internal healthy control values, Table 3). Conversely, inhibition of activity induced by antiplatelet acetylsalicylic acid (ASA) was found to be more pronounced on both post-COVID subjects as compared to the instrument normal instrument control.

**Table 3.** Whole blood impedance aggregometry

Whole blood impedance aggregometry
|  | No ASA |  |  |  | + ASA<sup>2</sup> |  |  |  |
| --- | --- | --- | --- | --- | --- | --- | --- | --- |
|  | Internal<br>healthy<br>control | Long-COVID<br>subject | Post-COVID<br>subject | Significance<sup>1</sup> | Internal<br>healthy<br>control | Long-COVID<br>subject | Post-COVID<br>subject | Significance<sup>1</sup> |
| Amplitude,<br>Ohm | 4.5 | 11.4 ±<br>0.2 | 8.6 ±<br>0.7 | $p < 0.05$ | 2.3 | 1.5 ±<br>0.2 | 1.1 ±<br>0.2 | ns |
| Rate,<br>Ohm/min | 1.5 | 8.1 ±<br>0.2 | 6.1 ±<br>0.2 | $p < 0.05$ | 0.5 | 0.45 ±<br>0.05 | 0.35 ±<br>0.05 | ns |
| Lag time,<br>min | 1.5 | 1.4 ±<br>0.1 | 1.3 ±<br>0.1 | ns | 3.5 | 2.3 ±<br>0.1 | 2.2 ±<br>0.1 | ns |
| AUC (at 10<br>min) | 35 | 80 ± 7 | 66 ± 5 | $p < 0.05$ | 15 | 9 ± 3 | 5 ± 2 | ns |

<sup>1</sup>Significance: T-test comparison between Long-COVID and asymptomatic post-COVID subjects;
<sup>2</sup>Supplemented with 100 µM aspirin
Note: Experimental error was evaluated based on signal to noise ratios of experimental data.

As compared to asymptomatic post-COVID subject, the Long-COVID subject further exhibited significantly elevated amplitude and AUC, and elevated maximum rate for the (Figure 5 and Table 3). Difference in lag time lacked significance. For both Long-COVID and asymptomatic post-COVID subjects, *in-vitro* ASA supplementation resulted in marked inhibition of aggregation with the resultant similar rates and lag times. Aggregation of platelets from both post-COVID asymptomatic and Long-COVID subjects, as indicated by aggregometry, was significantly inhibited by ASA with the differences in resultant aggregation lacking statistical significance (Table 3).

**Figure 5.**
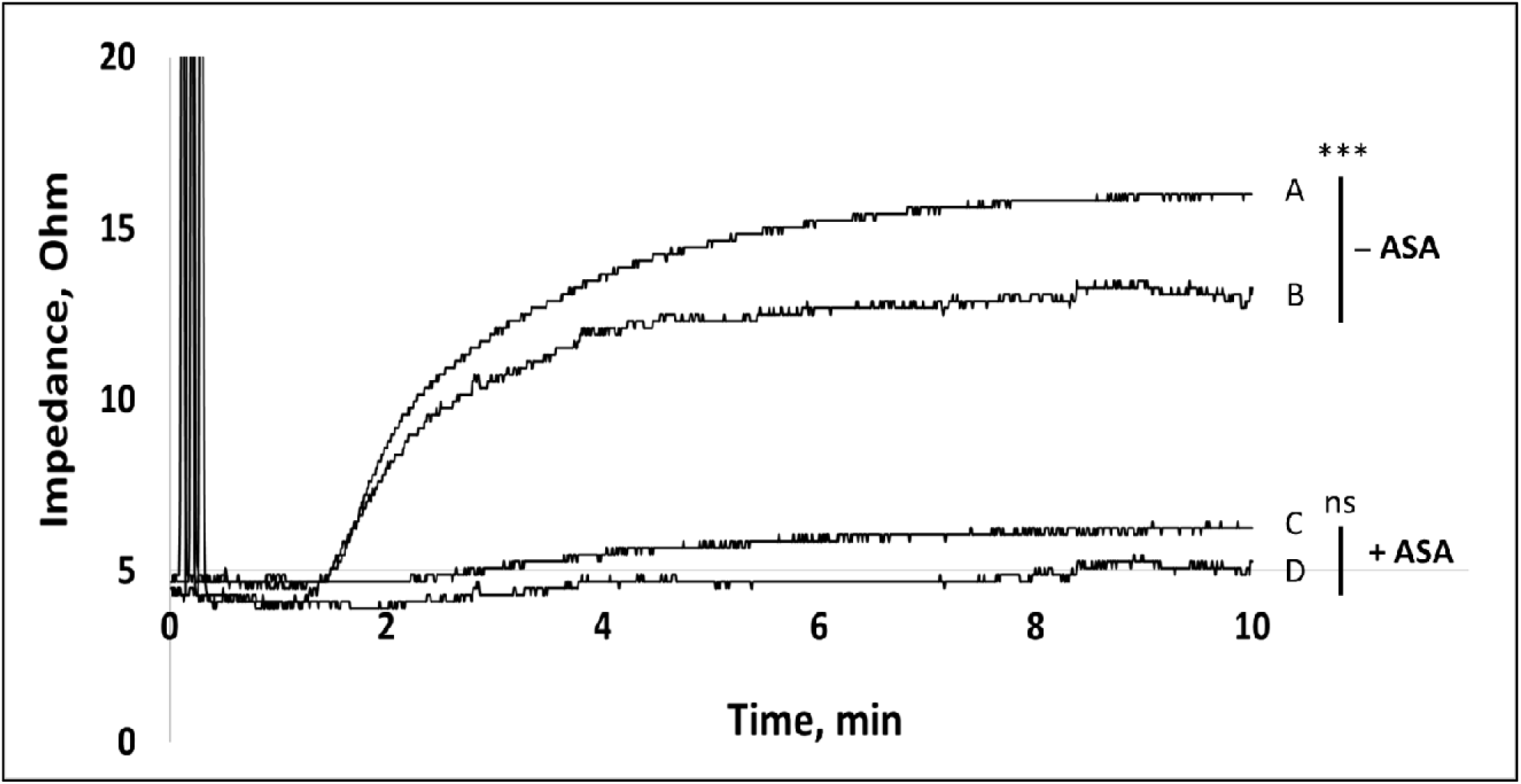
Whole blood impedance aggregometry for LC (Long-COVID subject, traces A and C) and PC (asymptomatic post-COVID, traces B and D) subjects with and without supplementation with 100 µM aspirin. *** indicates statistically significant difference at *p* < 0.05; ns – no significance.

## Discussion

It was demonstrated that platelets from COVID-19 patients overexpress P-Selectin both basally and upon activation, associated with faster platelet aggregation and increased circulating platelet– leukocyte aggregates [31]. Significantly higher levels of the platelet activation markers including P-Selectin, had also been reported for stable hospitalized COVID-19 patients [32]. Interestingly, no changes in platelet reactivity of P-selectin expression were detected upon vaccination within 4 weeks of BNT162b2 administration, with overall activation marker expression, including P-selectin, being significantly lower than even for mild-COVID-19 patient cohort [33]. While the drivers behind platelet activation in COVID-19 remain to be determined, it is possible, that such activation occurs due to platelet interaction with the infected endothelium and/or as a result of the cytokine storm associated with SARS-CoV-2 infection [34].

Overall, the evidence seems to point to the conclusion that SARS-CoV-2 infection is indeed associated with platelet hyperreactivity, contributing to COVID-19 pathophysiology [35]. A recently completed (results are pending) Phase 2 clinical trial to test Crizanlizumab, a P-Selectin inhibiting antibody, in hospitalized COVID-19 patients (ClinicalTrials.gov Identifier: NCT04435184) highlights the attention to the hypothesized role of P-Selectin in SARS-CoV-2 infection [36].

In this work we observe that whole blood *in-vitro* treatment with P-Selectin inhibitor (Crizanlizumab) at a clinically use dose while effective in reducing P-selectin cell adhesion in an asymptomatic post-COVID subject, was ineffective in the case of a patient with severe Long-COVID symptoms. Presented results are strongly indicative of the cause being linked to persistent platelet activation with P-Selectin overexpressed on platelets essentially “scrubbing” the drug from the environment. Thus, while no adhesion inhibition was observed in whole blood of the Long-COVID subject, crizanlizumab was effective in reducing adhesion to P-selectin when used with either i-WBC in buffer or when the assay was performed in autologous platelet poor plasma (PPP). Platelet activation suggested for Long-COVID patient (LC) by the flow adhesion study was further confirmed by significantly elevated platelet activity as shown by impedance aggregometry. Such activity we equally inhibited in the presence of ASA. As ASA inhibitory effect occurs through inhibition of cyclo-oxygenase, it would not be affected by levels of platelet P-Selectin expression [37].

Despite the mounting evidence detailing possible involvement of platelets in COVID-19, the cohesive picture of their involvement remains to be developed. Some investigations report increased platelet activation and platelet-monocyte aggregate formation in severe COVID-19 patients, but not in patients presenting mild COVID-19 syndrome [13], while other document platelet hyperactivity in COVID-19 independent of disease severity [16].

Ex vivo exposure of healthy donor platelets to plasma from severe COVID-19 patients may cause sustained control platelets activation, in terms of P-selectin expression and platelet–leukocyte aggregate formation, similar to that observed in vivo [38], and inducing tissue factor (TF) expression by the monocytes. Such TF expression was reduced by platelet pre-treatment with anti–P-Selectin neutralizing antibodies [13].

Platelet aggregation can be elevated in healthy women compared to men, with such sex-related difference present also after incubation with ASA [39]. A trend towards a higher platelet reactivity mainly associated with women exhibiting elevated (defined as >200 Billion cells/L) platelet counts was also reported by Ranucci et.al [40]. While gender-related factors cannot be conclusively excluded, the difference in platelet reactivity reported in this work is markedly larger than the 10% observed by Ivandic et.al [39], and likely does indicate a pathology-associated nature of the observed difference.

When tested in platelet poor plasma, adhesion on P-selectin was markedly decreased for both subjects, as compared to that in the buffer solution (see Figure 2). Such decline can be interpreted in terms of white cell PSGL-1 receptor binding with soluble P-selectin that would be present in subject’s plasma, but not in the buffer medium. Soluble P-selectin would be present in plasma in its monomeric, as opposed to oligomeric form typical for membrane bound P-selectin. However, only the lectin and epidermal growth factor domains are necessary for P-selectin interaction with PSGL-1 [41]. Thus, monomeric sP-selectin was shown to binds to high affinity ligands on leukocytes with its avidity for leukocytes enhanced when the protein is present in an oligomeric transmembrane form [42]. Significantly, the loss of surface P-selectin does not seem to prevent platelets from circulating and function, showing aggregation, adhesion, and procoagulant activity [21].

Higher activation status of platelets for the Long-COVID subject along with decreased efficacy of P-selectin adhesion inhibition described above, would seem to imply higher sP-selectin levels in Long-COVID autologous plasma [43]. However, sP-selectin plasma pool would also include sP-selectin originated from vascular endothelial cells and the significance of platelet contribution to it had not been assessed in this work. Evaluation of the role and significance of sP-selectin in mediation of whole blood and plasma response to P-Selectin inhibition therapy would be the subject of future research.

*In-vitro* treatment with crizanlizumab of i-WBC suspended in the autologous PPP resulted in comparable inhibition of flow adhesion (see Figure 4). However, in the same assay performed in buffer solution, i-WBC from the subject with severe Long-COVID symptoms showed much faster and stronger response, with close to complete inhibition of P-selectin adhesion even at sub-clinical crizanlizumab dose (see Figure 3). This seems to indicate either decreased PSGL-1 expression or more likely, a decrease in activation-dependent post-translational PSGL-1 modification. It is known that expression of the native protein is insufficient for selectin binding, and activation-associated post-translational modifications by sialic acid (SiA) and fucose are required to enable PSGL-1 binding to P-Selectin with high affinity [44]. Significantly, O-glycan modification involved in PSGL-1 activation was shown to regulate blood cell activity by influencing the balance between selectin-mediated cell rolling and cellular response to the chemotactic cues used in steady state T-cell traffic, reflecting platelet role as effector cell involved in both hemostasis and in pathological thrombosis [45].

Interestingly, SiA that is critically involved in PSGL-1 posttranslational modifications, is an established target for binding activity by many coronaviruses [46]. Comparing to influenza virus family which uses Hemagglutinin, a glycoprotein coat protein, to target SiA on host cell membranes, SARS-CoV-2, seems rely on more specific interactions facilitated by the virus S-protein. It is present on ACE2 receptors and is an integral part of, which present glycosphingolipid with one or more SiA molecule(s) linked to the sugar chain. On a cellular level, the entry pathway for SARS-CoV-2 was suggested to be through SiA-mediated interactions with the receptor-binding domain of the viral spike protein [47]. It was suggested that diverse clinical presentation of COVID-19 and specifically differences in age and gender associated severity may, in part, be explained through age and gender-related variability of sialome among the patients [48]. Overall, the clinical importance of SiA-mediated interactions between the host cell and virus certainly require further investigation [49].

SiA receptor-destroying enzymes like Neuraminidase (NEU) in influenza or hemagglutinin-esterase (HE) in some coronaviruses, mediate cleavage of surface SiA-containing cell receptors promoting viral particles release from infected cells. Endogenous cell NA activity plays a role in shedding of cell surface SiA, plays a key role in activation of both platelets [50] and neutrophils [51]. While SARS-CoV-2 seem to be lacking HE gene [52], samples from COVID-19 patients revealed an overexpression of NEU in infected neutrophils and treatment with NEU inhibitors led to decreased host NEU-mediated shedding of cell SiA and reduced overactivation of neutrophils [53]. The limitation of this discussion is that the possibility of leukocyte-platelet aggregates co-purifying during i-WBC isolation cannot be completely ruled out.

It remains uncertain whether SARS-CoV-2 is capable of established a persistent and stable chronic infection in human tissues like other SARS coronaviruses [54]. Nevertheless, viral RNA is known to persist in human body well past the resolution of the active infection [3] and SARS-CoV-2 RNA had been identified in samples from patients long time after the resolution of the active infection [55-57]. Considering that SARS-CoV-2 RNA present in the blood stream was associated with platelet hyperactivity [58, 59], a similar mechanism for platelet activation can be suggested for Long-COVID case. The key limitation of such a hypothesis would derive from the fact that the above studies observed SARS-CoV-2 RNA up to a maximum of 3 months after the infection, while in the presented case, the Long-COVID patient (LC) presented platelet hyperactivity at 20 weeks and experienced severe Long-COVID symptoms even at 28 weeks after the resolution of active COVID-19 disease. In that regard, the potential for persistent SARS-CoV-2 or presence of active viral RNA in patient body more than half a year post resolution had not been fully assessed. The uncertain potential for disease resurgence and/or spread gives this question extra importance and urgency.

The clear limitation of this work is the observation of a single Long-COVID patient, even if contrasted with an asymptomatic post-COVID subject. Moreover, it is not known to what extent *in-vitro* observations would translate to *in-vivo* treatment efficacy, especially to its long-term effects, with the treatment-related changes to cell function accumulating over time. Additionally, interaction of P-selectin inhibitors with overexpressed P-selectin on platelets of Long-COVID patients can potentially alter platelet function leading to possible clinical effects not directly observable through or related to cell adhesion to P-selectin. There could be significant potential differences in efficacy of P-selectin inhibitors drugs in terms of their action on the protein expressed on blood cells (likely, mostly platelets) as opposed to drugs’ impact on flow-mediated interaction with endothelially expressed immobilized P-selectin. However, relative significance of these two mechanisms of action to drug clinical efficacy still remains to be elucidated.

It also remains presently unknown to what extent the observed cell function alterations in severe Long-COVID case are indeed a general feature of the condition or a feature of clinical presentation in that particular patient. Clinical presentation reported for SARS-CoV-2 infection is highly variable, and such is likely to extend to Long-COVID sequalae. Individual variability could be expected to play a significant role, likely on a background of certain common condition-associated factors, potentially including platelet hyperactivity with P-Selectin overexpression and alteration in functionality of PSGL-1 leukocyte receptor reported here. Where such are general features of Long-COVID or a singular individual response, may be an important question affecting the scope of utility of P-selectin inhibitor drugs in Long-COVID treatment. Further work is warranted to expand our understanding on mechanisms defining altered cell functionality in Long-COVID.

## Conclusions

- Severe Long-COVID subject presented platelet hyperactivity associated with increased P-Selectin activity and seemingly with alteration in functionality of PSGL-1 leukocyte receptor, while the asymptomatic post-COVID subject did not present similar alterations of blood cell function.
- The data suggests significantly higher levels of p-selectin activity in Long-COVID, suggesting P-selectin activity as a potential driver of Long-COVID pathology.
- At similar concentrations, crizanlizumab, a p-selectin inhibitor, was significantly less effective in elucidating a response in Long-COVID patient as compared to asymptomatic post-COVID subject with several-fold larger dose was required to generate a moderate response in Long-COVID subject based on cell adhesion to immobilized P-selectin.

## Data Availability

All data produced in the present study are available upon reasonable request to the authors

## Acknowledgements

The authors gratefully acknowledge study participants, who contributed their time and blood for this study. Thank you!

## References

1. Datta SD, Talwar A, Lee JT. A Proposed Framework and Timeline of the Spectrum of Disease Due to SARS-CoV-2 Infection: Illness Beyond Acute Infection and Public Health Implications. Jama. 2020;324(22):2251–2.

2. Greenhalgh T, Knight M, A’Court C, Buxton M, Husain L. Management of post-acute covid-19 in primary care. BMJ (Clinical research ed). 2020;370:m3026.

3. Proal AD, VanElzakker MB. Long COVID or Post-acute Sequelae of COVID-19 (PASC): An Overview of Biological Factors That May Contribute to Persistent Symptoms. Frontiers in microbiology. 2021;12(1494).

4. Abrams JY, Godfred-Cato SE, Oster ME, Chow EJ, Koumans EH, Bryant B, et al. Multisystem Inflammatory Syndrome in Children Associated with Severe Acute Respiratory Syndrome Coronavirus 2: A Systematic Review. The Journal of pediatrics. 2020;226:45-54.e1.

5. Del Rio C, Collins LF, Malani P. Long-term Health Consequences of COVID-19. Jama. 2020;324(17):1723–4.

6. Puntmann VO, Carerj ML, Wieters I, Fahim M, Arendt C, Hoffmann J, et al. Outcomes of Cardiovascular Magnetic Resonance Imaging in Patients Recently Recovered From Coronavirus Disease 2019 (COVID-19). JAMA Cardiology. 2020;5(11):1265–73.

7. Carvalho-Schneider C, Laurent E, Lemaignen A, Beaufils E, Bourbao-Tournois C, Laribi S, et al. Follow-up of adults with noncritical COVID-19 two months after symptom onset. Clin Microbiol Infect. 2021;27(2):258–63.

8. Health F. Detailed Study of Patients with Long-Haul COVID: An Analysis of Private Healthcare Claims [White paper]2021. Available from: https://s3.amazonaws.com/media2.fairhealth.org/whitepaper/asset/A%20Detailed%20Study%20of%20Patients%20with%20Long-Haul%20COVID--An%20Analysis%20of%20Private%20Healthcare%20Claims--A%20FAIR%20Health%20White%20Paper.pdf.

9. Mardian Y, Kosasih H, Karyana M, Neal A, Lau C-Y. Review of Current COVID-19 Diagnostics and Opportunities for Further Development. Frontiers in medicine. 2021;8(562).

10. Deng X, Liu B, Li J, Zhang J, Zhao Y, Xu K. Blood biochemical characteristics of patients with coronavirus disease 2019 (COVID-19): a systemic review and meta-analysis. Clinical chemistry and laboratory medicine. 2020;58(8):1172–81.

11. Suklan J, Cheaveau J, Hill S, Urwin SG, Green K, Winter A, et al. Utility of Routine Laboratory Biomarkers to Detect COVID-19: A Systematic Review and Meta-Analysis. Viruses. 2021;13(5).

12. Samprathi M, Jayashree M. Biomarkers in COVID-19: An Up-To-Date Review. Frontiers in pediatrics. 2021;8(972).

13. Hottz ED, Azevedo-Quintanilha IG, Palhinha L, Teixeira L, Barreto EA, Pão CRR, et al. Platelet activation and platelet-monocyte aggregate formation trigger tissue factor expression in patients with severe COVID-19. Blood. 2020;136(11):1330–41.

14. Battinelli EM. COVID-19 concerns aggregate around platelets. Blood. 2020;136(11):1221–3.

15. Barrett TJ, Lee AH, Xia Y, Lin LH, Black M, Cotzia P, et al. Platelet and Vascular Biomarkers Associate With Thrombosis and Death in Coronavirus Disease. Circulation research. 2020;127(7):945–7.

16. Zaid Y, Puhm F, Allaeys I, Naya A, Oudghiri M, Khalki L, et al. Platelets Can Associate with SARS-Cov-2 RNA and Are Hyperactivated in COVID-19. Circulation research. 2020;127(11):1404–18.

17. Merten M, Thiagarajan P. P-selectin in arterial thrombosis. Zeitschrift fur Kardiologie. 2004;93(11):855–63.

18. Agrati C, Sacchi A, Tartaglia E, Vergori A, Gagliardini R, Scarabello A, et al. The Role of P-Selectin in COVID-19 Coagulopathy: An Updated Review. International journal of molecular sciences. 2021;22(15).

19. Agrati C, Bordoni V, Sacchi A, Petrosillo N, Nicastri E, Del Nonno F, et al. Elevated P-Selectin in Severe Covid-19: Considerations for Therapeutic Options. Mediterranean journal of hematology and infectious diseases. 2021;13(1):e2021016.

20. Fenyves BG, Mehta A, Kays KR, Beakes C, Margolin J, Goldberg MB, et al. Plasma P-selectin is an early marker of thromboembolism in COVID-19. American journal of hematology. 2021;96(12):E468–e71.

21. Michelson AD, Barnard MR, Hechtman HB, MacGregor H, Connolly RJ, Loscalzo J, et al. In vivo tracking of platelets: circulating degranulated platelets rapidly lose surface P-selectin but continue to circulate and function. Proceedings of the National Academy of Sciences of the United States of America. 1996;93(21):11877–82.

22. Panicker SR, Mehta-D’souza P, Zhang N, Klopocki AG, Shao B, McEver RP. Circulating soluble P-selectin must dimerize to promote inflammation and coagulation in mice. Blood. 2017;130(2):181–91.

23. Castanares-Zapatero D, Chalon P, van den HEEDE K. Pathophysiology of Long-COVID: a Preliminary Report. Federaal Kenniscentrum voor de Gezondheidzorg; 2021.

24. Hines PC, Callaghan MU, Zaidi AU, Gao X, Liu K, White J, et al. Flow adhesion of whole blood to P-selectin: a prognostic biomarker for vaso-occlusive crisis in sickle cell disease. British journal of haematology. 2021;194(6):1074–82.

25. Kapileshwarkar Y, Gelmini L, Tseng Y-S, Jackson T, Gao X, Richards K, et al. Assessment of antiplatelet therapy response in pediatric patients following cardiac surgery by microfluidic assay. Progress in Pediatric Cardiology. 2020;56:101191.

26. McShine RL, Sibinga S, Brozovic B. Differences between the effects of EDTA and citrate anticoagulants on platelet count and mean platelet volume. Clinical and laboratory haematology. 1990;12(3):277–85.

27. Mehta AA, Haridas N, Belgundi P, Jose WM. A systematic review of clinical and laboratory parameters associated with increased severity among COVID-19 patients. Diabetes & metabolic syndrome. 2021;15(2):535–41.

28. Rostami M, Mansouritorghabeh H. D-dimer level in COVID-19 infection: a systematic review. Expert review of hematology. 2020;13(11):1265–75.

29. Malka R, Delgado FF, Manalis SR, Higgins JM. In vivo volume and hemoglobin dynamics of human red blood cells. PLoS computational biology. 2014;10(10):e1003839.

30. Ataga KI, Kutlar A, Kanter J, Liles D, Cancado R, Friedrisch J, et al. Crizanlizumab for the Prevention of Pain Crises in Sickle Cell Disease. The New England journal of medicine. 2017;376(5):429–39.

31. Manne BK, Denorme F, Middleton EA, Portier I, Rowley JW, Stubben C, et al. Platelet gene expression and function in patients with COVID-19. Blood. 2020;136(11):1317–29.

32. Bongiovanni D, Klug M, Lazareva O, Weidlich S, Biasi M, Ursu S, et al. SARS-CoV-2 infection is associated with a pro-thrombotic platelet phenotype. Cell death & disease. 2021;12(1):50.

33. Klug M, Lazareva O, Kirmes K, Rosenbaum M, Lukas M, Weidlich S, et al. Platelet expression and reactivity after BNT162b2 vaccine administration. medRxiv : the preprint server for health sciences. 2021:2021.05.18.21257324.

34. Bikdeli B, Madhavan MV, Jimenez D, Chuich T, Dreyfus I, Driggin E, et al. COVID-19 and Thrombotic or Thromboembolic Disease: Implications for Prevention, Antithrombotic Therapy, and Follow-Up: JACC State-of-the-Art Review. Journal of the American College of Cardiology. 2020;75(23):2950–73.

35. Comer SP, Cullivan S, Szklanna PB, Weiss L, Cullen S, Kelliher S, et al. COVID-19 induces a hyperactive phenotype in circulating platelets. PLoS biology. 2021;19(2):e3001109.

36. Neri T, Nieri D, Celi A. P-selectin blockade in COVID-19-related ARDS. American journal of physiology Lung cellular and molecular physiology. 2020;318(6):L1237–l8.

37. Taylor ML, Ilton MK, Misso NL, Watkins DN, Hung J, Thompson PJ. The effect of aspirin on thrombin stimulated platelet adhesion receptor expression and the role of neutrophils. British journal of clinical pharmacology. 1998;46(2):139–45.

38. Canzano P, Brambilla M, Porro B, Cosentino N, Tortorici E, Vicini S, et al. Platelet and Endothelial Activation as Potential Mechanisms Behind the Thrombotic Complications of COVID-19 Patients. JACC: Basic to Translational Science. 2021;6(3):202–18.

39. Ivandic BT, Giannitsis E, Schlick P, Staritz P, Katus HA, Hohlfeld T. Determination of aspirin responsiveness by use of whole blood platelet aggregometry. Clinical chemistry. 2007;53(4):614–9.

40. Ranucci M, Aloisio T, Di Dedda U, Menicanti L, de Vincentiis C, Baryshnikova E, et al. Gender-based differences in platelet function and platelet reactivity to P2Y12 inhibitors. PloS one. 2019;14(11):e0225771.

41. Mehta P, Patel KD, Laue TM, Erickson HP, McEver RP. Soluble Monomeric P-Selectin Containing Only the Lectin and Epidermal Growth Factor Domains Binds to P-Selectin Glycoprotein Ligand-1 on Leukocytes. Blood. 1997;90(6):2381–9.

42. Ushiyama S, Laue TM, Moore KL, Erickson HP, McEver RP. Structural and functional characterization of monomeric soluble P-selectin and comparison with membrane P-selectin. Journal of Biological Chemistry. 1993;268(20):15229–37.

43. Yatim N, Boussier J, Chocron R, Hadjadj J, Philippe A, Gendron N, et al. Platelet activation in critically ill COVID-19 patients. Annals of intensive care. 2021;11(1):113.

44. Vachino G, Chang XJ, Veldman GM, Kumar R, Sako D, Fouser LA, et al. P-selectin glycoprotein ligand-1 is the major counter-receptor for P-selectin on stimulated T cells and is widely distributed in non-functional form on many lymphocytic cells. The Journal of biological chemistry. 1995;270(37):21966–74.

45. Carlow DA, Gossens K, Naus S, Veerman KM, Seo W, Ziltener HJ. PSGL-1 function in immunity and steady state homeostasis. Immunological reviews. 2009;230(1):75–96.

46. Matrosovich M, Herrler G, Klenk HD. Sialic Acid Receptors of Viruses. Topics in current chemistry. 2015;367:1–28.

47. Sun XL. The Role of Cell Surface Sialic Acids for SARS-CoV-2 Infection. Glycobiology. 2021.

48. Morniroli D, Giannì ML, Consales A, Pietrasanta C, Mosca F. Human Sialome and Coronavirus Disease-2019 (COVID-19) Pandemic: An Understated Correlation? Frontiers in immunology. 2020;11(1480).

49. Dhar C, Sasmal A, Diaz S, Verhagen A, Yu H, Li W, et al. Are sialic acids involved in COVID-19 pathogenesis? Glycobiology. 2021;31(9):1068–71.

50. Lauková L, Weiss R, Semak V, Weber V. Desialylation of platelet surface glycans enhances platelet adhesion to adsorbent polymers for lipoprotein apheresis. The International journal of artificial organs. 2021;44(6):378–84.

51. Cross AS, Wright DG. Mobilization of sialidase from intracellular stores to the surface of human neutrophils and its role in stimulated adhesion responses of these cells. The Journal of clinical investigation. 1991;88(6):2067–76.

52. Zandi M, Behboudi E, Soltani S. Role of Glycoprotein Hemagglutinin-Esterase in COVID-19 Pathophysiology? Stem cell reviews and reports. 2021:1–2.

53. Formiga RO, Amaral FC, Souza CF, Mendes D, Wanderley CWS, Lorenzini CB, et al. Neuraminidase inhibitors rewire neutrophil function in vivo in murine sepsis and ex vivo in COVID-19. bioRxiv. 2021.

54. Chan PK, To KF, Lo AW, Cheung JL, Chu I, Au FW, et al. Persistent infection of SARS coronavirus in colonic cells in vitro. J Med Virol. 2004;74(1):1–7.

55. Liotti FM, Menchinelli G, Marchetti S, Posteraro B, Landi F, Sanguinetti M, et al. Assessment of SARS-CoV-2 RNA Test Results Among Patients Who Recovered From COVID-19 With Prior Negative Results. JAMA Internal Medicine. 2021;181(5):702–4.

56. Vibholm LK, Nielsen SSF, Pahus MH, Frattari GS, Olesen R, Andersen R, et al. SARS-CoV-2 persistence is associated with antigen-specific CD8 T-cell responses. EBioMedicine. 2021;64:103230.

57. Sun J, Xiao J, Sun R, Tang X, Liang C, Lin H, et al. Prolonged Persistence of SARS-CoV-2 RNA in Body Fluids. Emerging infectious diseases. 2020;26(8):1834–8.

58. Zhang S, Liu Y, Wang X, Yang L, Li H, Wang Y, et al. SARS-CoV-2 binds platelet ACE2 to enhance thrombosis in COVID-19. Journal of hematology & oncology. 2020;13(1):120.

59. Zaid Y, Puhm F, Allaeys I, Naya A, Oudghiri M, Khalki L, et al. Platelets Can Associate With SARS-CoV-2 RNA and Are Hyperactivated in COVID-19. Circulation research. 2020;127(11):1404–18.

